# Research Mentorship in Low and Middle-Income Countries: A Global Qualitative Evidence Synthesis of Data from a Crowdsourcing Open Call and Scoping Review

**DOI:** 10.1101/2022.09.19.22280121

**Authors:** Eneyi E. Kpokiri, Kamryn McDonald, Joseph A. Gebreyohannes, Lyda Osorio, Tilak Chandra Nath, Victor A. Talavera-Urdanivia, Pheabian O. Akinwale, Yuka C. Manabe, Barbara Castelnuovu, Weiming Tang, Daniel Yilma, Michael Mihut, Oliver Ezechi, Juliet Iwelunmor, Mirgissa Kaba, Alemseged Abdissa, Joseph D. Tucker

## Abstract

**Introduction:** Research mentorship is critical for advancing science, but there are few practical strategies for cultivating research mentorship in resource-limited settings. WHO/TDR Global commissioned a group to develop a practical guide on research mentorship. This global qualitative evidence synthesis included data from a crowdsourcing open call and scoping review to identify strategies to enhance research mentorship in low- and middle-income country (LMIC) institutions.

**Methods:** The crowdsourcing open call used methods recommended by WHO/TDR and solicited descriptions of strategies to enhance research mentorship in LMICs. The scoping review used the Cochrane Handbook and pre-defined the approach in a protocol. We extracted studies focused on enhancing research mentorship in LMICs. Textual data describing research mentorship strategies from the open call and studies from the scoping review were coded into themes. The quality of evidence supporting themes was assessed using the CERQUAL approach.

**Results:** The open call solicited 123 practical strategies and the scoping review identified 73 studies. We identified three broad trends related to engaging institutions across the life cycle of research mentorship, leveraging existing resources for research and training to expand research mentorship, and strengthening monitoring and evaluation of research mentorship programs. We identified the following strategies to enhance research mentorship: recognizing mentorship as an institutional responsibility that should be provided and expected from all team members (8 strategies, 15 studies; moderate confidence); leveraging existing research and training resources to enhance research mentorship (15 strategies, 49 studies; moderate confidence); digital tools to match mentors and mentees and sustain mentorship relations over time (14 strategies, 11 studies; low confidence); nurturing a culture of generosity so that people who receive mentorship then become mentors to others (7 strategies, 7 studies; low confidence); peer mentorship defined as informal and formal support from one researcher to another who is at a similar career stage (16 strategies, 12 studies; low confidence).

**Interpretation:** Research mentorship can be strengthened in resource-limited institutions. The evidence from this open call and scoping review informed a WHO/TDR practical guide. More research mentorship programs are needed in LMIC institutions.

## Introduction

Mentorship is fundamental to global health research.^1^ Mentorship is often catalytic in launching individual research careers, building research teams at the group level, and sustaining research institutions over time.^2^ Although the individual and institutional pillars of research mentorship are widely recognized, there are fewer resources focused on enhancing research mentorship at the institutional level.^3^ Institutions (e.g., universities, research institutes, and other groups) create mentorship expectations, programs, incentives, and policies.

Research mentorship tools have been mainly designed for high-income research institutions, neglecting low and middle-income countries (LMICs).^4^ LMIC institutions may have different traditions, structures, cultures, and capacities related to research mentorship. For example, LMIC institutions often have fewer training grants focused on building research mentorship, comparatively fewer senior mentors per mentee, and less institutional support. At the same time, there are many indigenous research mentorship strategies that suggest LMIC-centered approaches are feasible and effective.^2^

In response, we organized a crowdsourcing open call and scoping review to identify strategies to enhance research mentorship in LMICs. A crowdsourcing open call has a group of people solve a problem and then implement selected solutions.^5^ The open call and scoping review were commissioned by the UNICEF/UNDP/World Bank/WHO Special Programme for Research and Training in Tropical Diseases, TDR. TDR Global is a global group of scientists and experts passionate about building capacity for research on neglected infectious diseases.

This global qualitative evidence synthesis included data from the crowdsourcing open call and scoping review in order to identify strategies to enhance research mentorship in LMIC institutions.

## Methods

### Crowdsourcing open call

The crowdsourcing open call on research mentorship was organized by Social Entrepreneurship to Spur Health (SESH), TDR Global, and the Armauer Hansen Research Institute (AHRI). The open call was implemented using stages outlined in the in the TDR/SESH/SIHI practical guide on crowdsourcing.^6,7^ The open call was launched on 21^st^ October 2021 and closed on 15^th^ February 2022. The open call had several steps including (1) organized a multisectoral steering committee; (2) engaged the community to contribute; 3) independently evaluated submissions; (4) recognized finalist participants; (5) refined ideas and implemented selected ideas (Table 1).

**Table 1.**
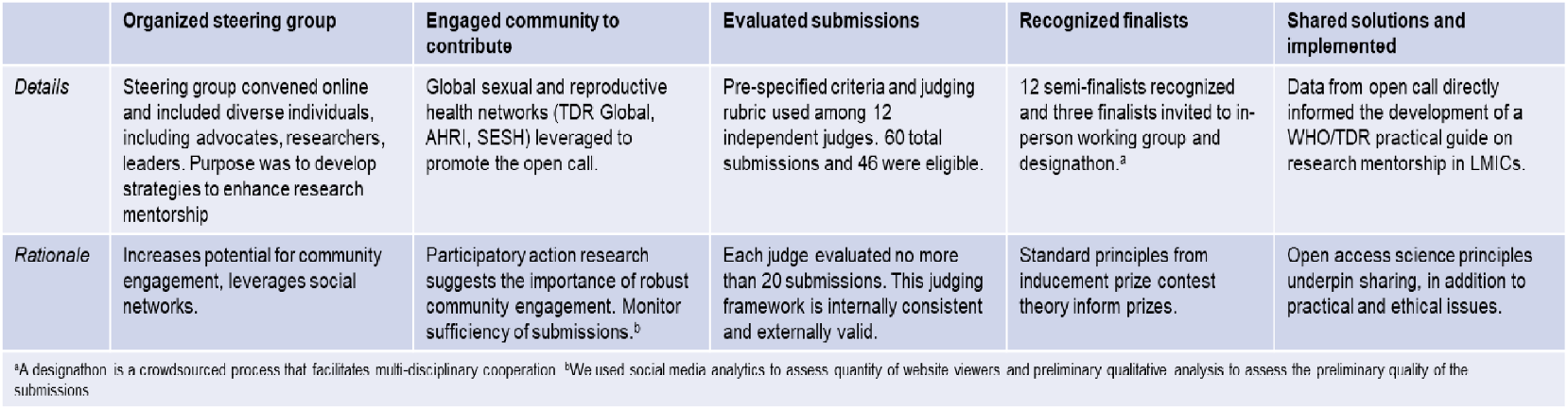
Outline of the main steps of the crowdsourcing open call. Developed based on the TDR/SIHI/SESH Practical Guide on crowdsourcing in Health and Health Research and the SIHI crowdsourcing Open Call Guidelines.

We organized a multisectoral steering committee from key stakeholders across diverse geographic regions. Steering committee members were recruited based on previous experience in research mentorship projects, their country of residence (prioritizing LMIC researchers), and their previous experience with TDR Global. The 25-person steering committee members included public health experts, government leaders, training directors, social media and communications experts, and clinical physicians.

The open call was announced on the website, social media, and through partner organization networks. Participants were invited to submit ideas for strategies that strengthened existing initiatives, established new mentorship programs, and aimed to create and sustain strong cultures of research mentorship. The website included a translation widget to facilitate non-English speaker participation. The website included the purpose of the open call, background information, judging criteria, eligibility criteria, formatting details, and deadlines. All participants were also asked to complete a brief online survey that gathered demographic data including age, gender, country of residence, and education level.

The open call was promoted through digital networks including social media (Twitter, Facebook, and LinkedIn), email listservs, and networks of collaborating organizations represented by the steering committee. Corresponding authors of studies in the scoping review were contacted to identify similar evidence. We used real-time social media analytics from the website, Twitter activity, and submissions to assess regional engagement and re-direct promotion efforts. All promotional materials, social media cards, and emails were translated into Spanish and French.

A total of twelve independent judges rated each submission 1-10 scale (1 as low and 10 as high) in five categories: 1) clear description; 2) potential for enhancing research mentorship in LMICs; 3) innovation; 4) potential for transferability in diverse LMIC settings; 5) promotion of equity and fairness. In addition, each judge gave an overall score of 5-50. Judges with a conflict of interest recused themselves from reviewing that entry. Conflicts were defined as collaborating, co-authoring, helping, receiving, or providing monetary or other support, or anything that could be perceived as a conflict of interest. The non-English entries were evaluated for initial eligibility with the use of a translation software and then were judged by those proficient in the language of the entry. An overall mean score that averaged the means for each of the five subcomponents (clear description, potential for enhancing research mentorship in LMICs, innovation, potential for transferability in diverse LMIC settings, promotion of equity and fairness) was calculated.

A mean score of greater than 35 (a threshold pre-specified by the Steering Committee,) on the 5-50 scale received a commendation of excellence on behalf of TDR Global, SESH and AHRI. Five finalists were ultimately selected based on the judging criteria. The five open call finalists were invited to join a WHO/TDR virtual working group to contribute to the development of a WHO/TDR practical guide. Three finalists were also invited to attend a TDR Global/AHRI conference on research mentorship in Addis Ababa, Ethiopia on June 23rd to June 24th, 2022.

### Scoping Review

A concurrent scoping review was conducted using the Preferred Reporting Items for Systematic Reviews and Meta-Analyses extension for Scoping Reviews (PRISMAScR).^8^ The purpose was to summarize available practices, lessons, and gaps/needs of health-related research mentorship programs in LMICs. Keyword search was conducted in PubMed, EMBASE, Cochrane Database of Systematic Reviews, JBI Evidence Synthesis, EBSCO, SciELO (Scientific Electronic Library Online) and AJOL (African Journals Online). Grey literature was also searched using included ProQuest Dissertations and Theses, Google Scholar, and institutional websites (National Institute of Health). The search results were screened using a pre-defined set of inclusion and exclusion criteria, and eligible articles for inclusion in the review were collated for data extraction. Inclusion criteria were focused on research mentorship and implemented in LMICs.

### Data Analysis

From the open call submissions, we received both quantitative data and qualitative data and we used a parallel mixed methods approach to analyze and present the data.^9^ The quantitative data, included the participants demographics and submission characteristics. This data was analyzed and presented using basic descriptive frequencies. The data from included articles and the content of the submissions were mostly textual qualitative data, and this was analyzed thematically using the framework approach including familiarization with the data, coding, charting and mapping out themes for interpretation.^10,11^ The themes identified from the open call were further strengthened with findings from the scoping review (Figure 1). To ensure validity and reliability in presenting findings, the eligible submissions and included articles were coded separately by two independent reviewers (EK and KM) and discrepancies were reviewed by a third team member (JDT). We used the GRADE-CERQual (Confidence in the Evidence from Reviews of Qualitative research) approach to assess confidence in the certainty of the review findings.^12^ This includes an assessment based on methodological rigor, coherence of the review finding, adequacy of the data, and relevance of the included studies to the review question.^13-18^ Each of these components was assessed and made into an overall judgement on the confidence in each review finding (very low, low, moderate, high).

**Figure 1:**
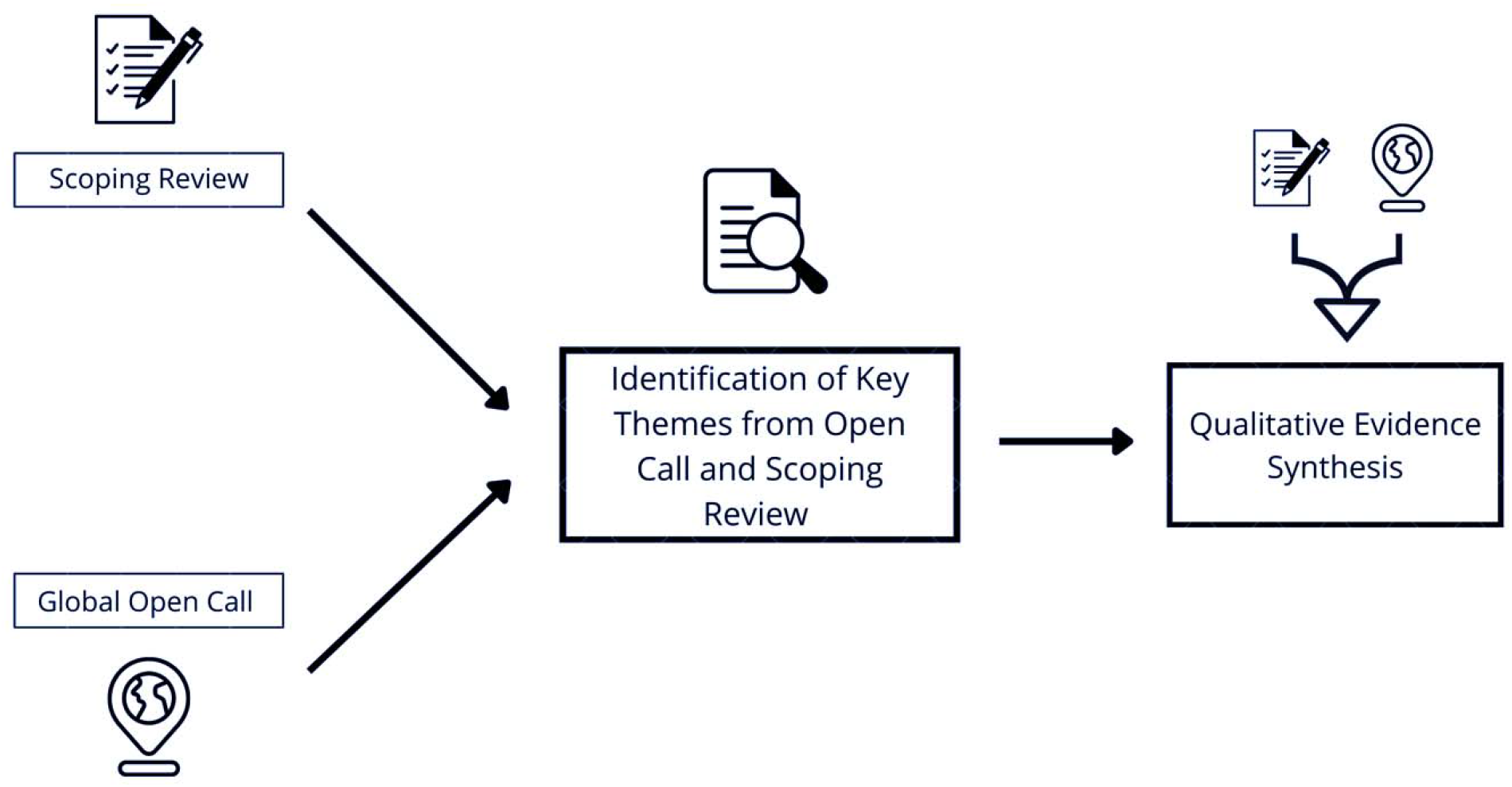
Stages in the identification of key themes.

### Ethical Considerations

The final compilation of data to inform a practical guide and qualitative evidence synthesis was approved by the AHRI (PO/23/22) and London School of Hygiene and Tropical Medicine institutional review boards. The scoping review was also registered on the Open Science Framework platform (10.17605/OSF.IO/JQA9Z).

## Results

The open call received a total of 60 submissions and 46 submissions were eligible for judging. Twelve submissions met the pre-specified criteria for excellence (Figure 2).

**Figure 2:**
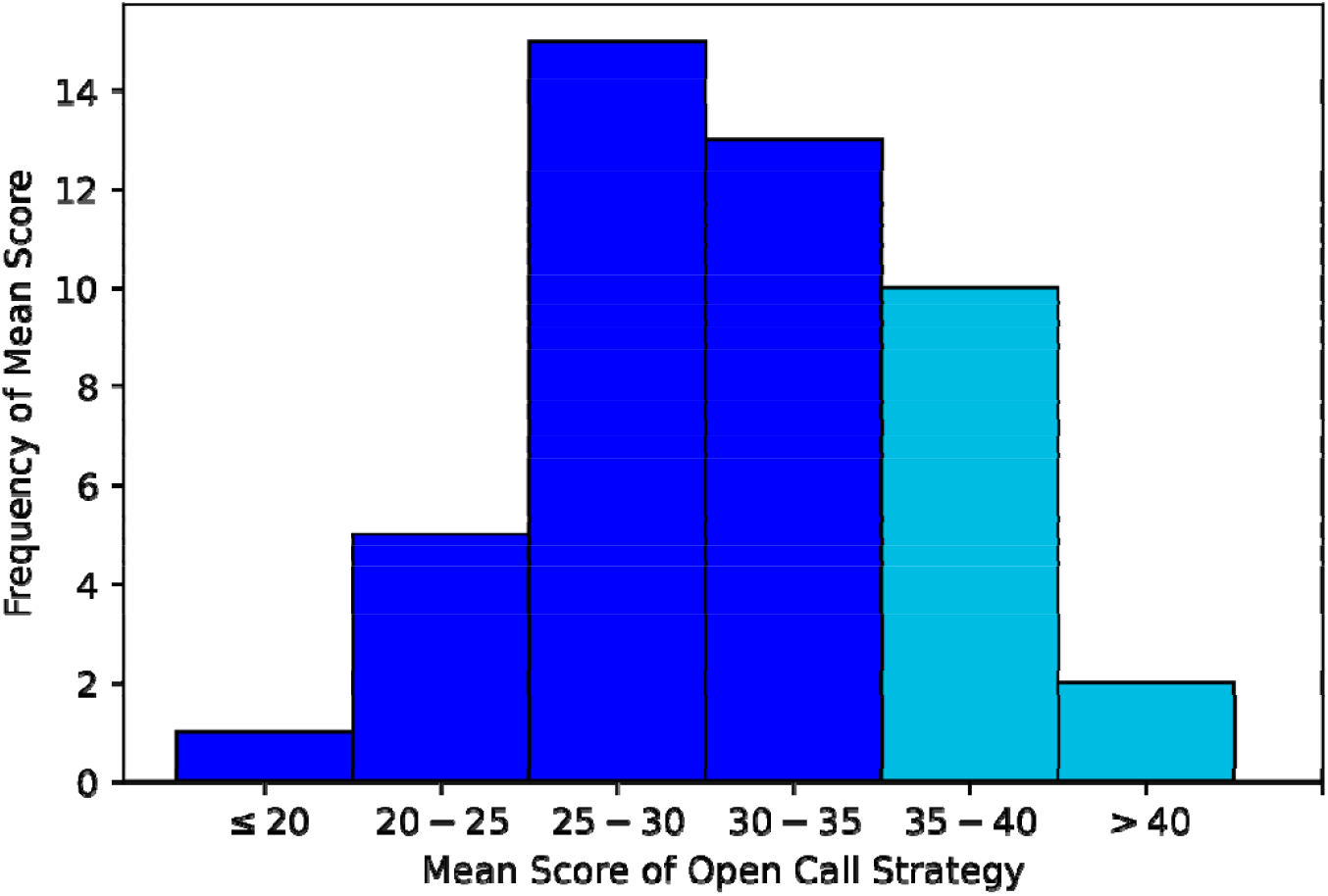
Distribution of mean scores on a 5-50 scale.

### Submission Characteristics and Participant demographics

We received submissions from a total of 33 different countries including 24 submissions from low-income countries, 18 submissions from middle-income countries, and four submissions from high-income countries. Top countries include Nigeria (10 submissions), Ethiopia (six submissions), Malawi (three submissions). Of the eligible submissions, 40 were in English language and six were submitted in Spanish. Table 2 below summarises the characteristics of submissions received.

**Table 2:** Characteristics of global research mentorship open call participants 2021-2022 (N = 45).

| **Characteristics** | **All eligible participants**<br>**(N=45)** |
| --- | --- |
| **Region** |  |
| Europe | 2 |
| Latin America | 6 |
| South Asia | 2 |
| Sub-Saharan Africa | 28 |
| East Africa | 4 |
| Middle East and North Africa | 1 |
| North America | 2 |
| **Country Economic Status** |  |
| HIC | 4 |
| MIC | 18 |
| LIC | 24 |
| **Gender** |  |
| Male | 24 |
| Female | 21 |
| **Degree** |  |
| High School Diploma | 3 |
| Bachelor's Degree | 7 |
| Masters Similar Professional Degree | 13 |
| Doctoral Degree | 22 |
| **Age** |  |
| 18-27 years old | 7 |
| 28-37 years old | 16 |
| 38-47 years old | 15 |
| 48 years old or older | 7 |

We had submissions from 26 male and 20 female participants. In terms of age, 16 applicants are 38-47 years old; 16 applicants are 28-37 years old; 7 applicants are 48 years old or older and 7 applicants are 18-27 years old. Most applicants had doctoral degrees (27 applicants), followed by applicants with master’s degrees (17 applicants), undergraduate degrees (one applicant) and high school degree (one applicant).

### Strategies to enhance mentorship

Themes identified from the analysis of the open call submissions have been broadly categorized into strategies that can enhance research mentorship within LMIC institutions. A total of 10 themes were identified from the results. The working group met and examined the evidence supporting each theme and five of these were prioritized as key. Key findings include recognizing mentorship as an institutional responsibility, leveraging existing research and training resources, digital tools to kickstart and sustain mentorship relations over time, nurturing a culture of generosity and peer mentorship (Figure 3).

**Figure 3:**
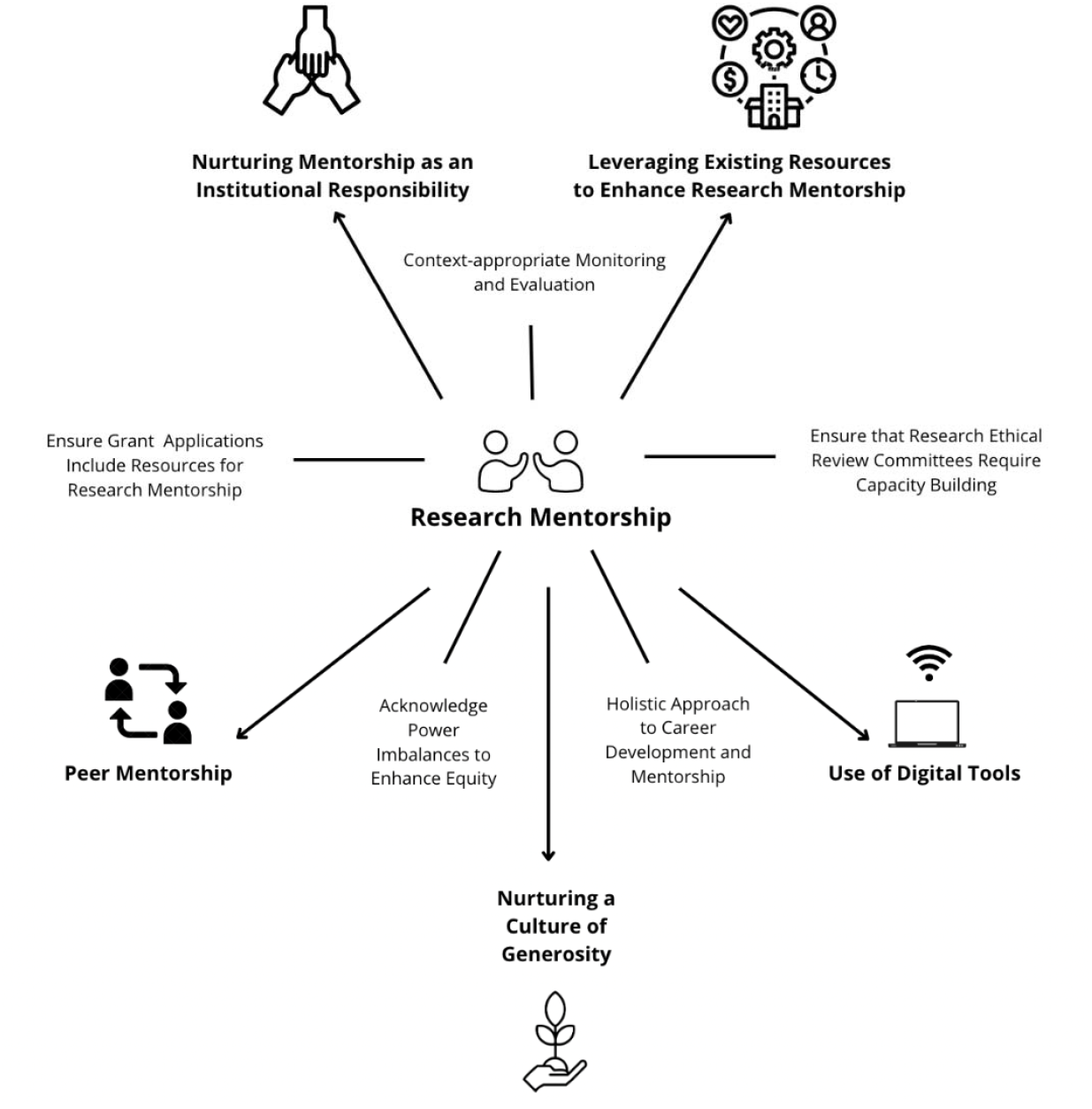
Overview of strategies to enhance research mentorship in LMICs (Top five themes in bold fonts)

#### Mentorship as an institutional responsibility

Eight strategies and 15 studies ^19-33^ identified research mentorship as an institutional and collective responsibility which should be expected from and provided by all team members. To support this, some submissions highlighted the development of a quick reference guide or policies to ensure that everyone is engaged in mentorship (5 submissions). Specific strategies to ensure institution-wide coverage include building research mentorship training into routine onboarding procedures, requiring research ethics committees to consider mentorship, and requiring grant applications to support research mentorship.

#### Digital tools to support mentorship

A total of 14 strategies and 11 studies^27,31,34-42^ demonstrated that digital tools enhanced research mentorship in LMICs. Digital tools including apps, websites, and other web-based platforms to aid in mentor/mentee matching, communication, establish and sustain mentorship relationship over time (14 submissions). While some strategies required internet access and sufficient bandwidth, there were many low-tech solutions that would be relevant in resource-constrained settings. These included mobile instant text messaging apps, social media groups such as Facebook and WhatsApp. However, several studies mentioned that digital approaches may exacerbate inequalities (eight studies) and be less relevant in a large number of LMIC institutions with limited digital infrastructure.^43-46, 47-50^ Additional challenges with digital tools in LMICs include power cuts and constant interruptions to internet access.^51^

#### Leveraging existing research and training resources

Fifteen open call strategies and 49 studies^19,20,22-32,35,37,38,40,42,52-83^ showed that leveraging existing research and training programs facilitated research mentorship. Already existing training programs with databases of staff, contacts/institutions that can be partnered with to network on mentorship and leverage other institution’s resources is beneficial for facilitating sustainability research mentorship. Leveraging established relationships with other institutions to create inter-institutional mentor/mentee relationships (6 submissions). Another example is the use of formal research project supervision with mentors acting as supervisors and mentees acting as researchers (7 submissions).

#### Culture of generosity

Seven open call strategies and seven studies^23,26,29-31,74,84^ included in the review emphasized the need for a culture of generosity and wholistic approach in mentorship relationships. Mentors are to consider other social determinants of life, be respectful of diversity and protect mentees from negative critics and racial bias. This warm and inclusive culture of generosity is thought to facilitate and develop motivation in mentees to become future mentors. Mentorship should purposely cultivate a culture of cooperation rather than competition within mentor/mentee programs (5 submissions).

#### Peer and group mentoring

A total of 16 open call submissions and 12 studies^23,27,29-31,40,76,80,84-87^ recognized the role of peer mentorship in enhancing research mentorship in LMIC settings. Peer mentorship is defined as informal and formal support from one researcher to another who is at a similar career stage. Introducing junior scientists and PhD’s into mentoring early on to create a culture of mentorship that is sustainable over time (4 submissions). Creation of a formal or organized mentorship club, group, or other form of community to aid in creating lasting mentor/mentee relationships (14 submissions). However, two studies highlighted hesitancy and barriers in peer mentorship that include fear, embarrassment, lack of knowledge and awareness.^47,48^

A summary of these key findings, the contributions strategies and studies and assessment of the certainty of the evidence of the finding are presented in the GRADE CERQual table below (Table 3).

**Table 3:** Evidence profile and assessment of confidence in the review findings as per GRADE-CERQual methodology.

| Study finding | Studies/ strategies contributing to the finding | Methodological limitations | Coherence | Adequacy | Relevance | CERQual Assessment |
| --- | --- | --- | --- | --- | --- | --- |
| **Institutional responsibility:** Recognize research mentorship as an institutional and collective responsibility to be provided by and expected from all team members. | OC=8 strategies<br>SR=15 studies | Minor concerns | Minor concerns | Serious concerns | Minor concerns | Moderate confidence |
| **Digital tools:** Digital tools can enhance institutionalization of research mentorship in LMICs by establishing and sustaining mentorship relationship over time | OC=14 strategies<br>SR=11 studies | Moderate concerns | Moderate concerns | Serious Concerns | Minor concerns | Low confidence |
| **Leveraging existing research and training programs:** Existing research and training programs facilitates institutional research mentorship | OC=15 strategies<br>SR=49 studies | No or minor concerns | Minor concerns | Minor concerns | No or Minor concerns | Moderate confidence |
| **Culture of generosity:** Mentors practicing warm and inclusive culture of generosity motivates mentees to become future mentors. | OC= 7 strategies<br>SR=7 studies | Minor concerns | Moderate concerns | Moderate concerns | Minor concerns | Low confidence |
| **Peer and group mentorship:** Informal and formal support from one researcher to another who is at a similar career stage enhances mentorship cultures in LMIC institutions. | OC=16 strategies<br>SR=12 studies | Moderate concerns | Serious concerns | Serous concerns | Minor concerns | Low confidence |

*OC- Open call* *SR- Scoping review*

## Discussion

Our data provide evidence-based strategies to improve research mentorship at LMIC institutions. Institutions need to recognize that research mentorship is a collective responsibility that should be expected and provided to all members. Promoting a culture of generosity increases the sense of collective responsibility for research mentorship. Ongoing research and training resources can be leveraged to spur research mentorship at the institutional level. This manuscript extends the literature^2^ by centering evidence and strategies from LMIC researchers, including data from a global crowdsourcing open call, and assessing the strength of the evidence using the CERQUAL approach.

Our data suggest that institutions should be responsible for ensuring that research mentorship is provided to and expected from all members. This contrasts the practice of many LMIC research mentorship programs that are offered to a subset of people.^88,89^ There are several strong reasons to consider research mentorship as a fundamental right of being in a university or research institute. Research mentorship can enhance recruitment and retention of promising research talents, build a sense of common purpose, and enhance research outcomes. Positive mentorship experiences have been correlated with greater institutional support for mentorship.^90^ Unfortunately, under-represented racial/ethnic minorities less often have research mentors compared to other researchers.^91,92^ Policies that make research mentorship available to all members could decrease mentoring disparities.

Our data suggest that nurturing a culture of generosity within research institutions can increase the likelihood of current mentees becoming subsequent mentors. This is consistent with research showing that people who receive mentorship are more likely to serve as mentors for other people.^93^ Mentorship within research institutions could create virtuous cycles that spur further kindness between researchers. Positive mentorship experiences provide examples of behaviors to emulate; negative mentorship experiences could be useful as a reminder of what not to do when you are a mentor.^93^

LMIC institutions have research and training resources that can be leveraged to enhance research mentorship. This finding aligns with previous mentoring toolkits^94^ and mentorship guidance.^1^ From a research perspective, obtaining research grants, joining professional associations, participating in conferences, and contributing to academic journals can expand opportunities for research mentorship. From a training perspective, using open access learning materials (e.g., massive open online courses), developing peer mentorship groups, and organizing university elective credit for research can formalize research mentorship.

This study has limitations. First, although, we received fewer non-English submissions and our relatively English-focused promotion materials likely limited contributions from some LMICs. At the same time, we accepted submissions in each of the five official United Nations languages and translated the call for submissions. Second, research mentorship is a complex and nuanced topic. Anticipating this complexity, we decided to accept text and non-text submissions. Third, published literature on research mentorship in LMICs is limited. Our use of a global crowdsourcing open call allowed us to elevate the voices of LMIC researchers and learn from indigenous strategies that have not yet been peer reviewed.

The data from this qualitative evidence synthesis have implications for research mentorship programs and policy. From a program perspective, these data suggest that research mentorship programs should be embedded within institutions and provided to all researchers. Expanding the scope of research mentorship could help decrease disparities in mentoring and build a sense of collective solidarity. From a policy perspective, the data from this qualitative evidence synthesis directly informed a WHO/TDR practical guide called HERMES, HEalth Research MEntorship in Low and Middle-Income CountrieS.

Research mentorship is a critical component of developing vibrant research institutions in LMICs. The evidence identified through this global open call and scoping review provide specific strategies and guiding principles for research mentorship. Research on implementation strategies to enhance mentorship at LMIC institutions is needed to advance this field. Monitoring and evaluation of research mentorship are critical for sustained success.

## Data Availability

All data produced in the present study are available upon reasonable request to the authors

## Acknowledgements

We would like to thank Annabel Steiner for extracting data on included studies. This research was supported by the UNICEF/UNDP/World Bank/WHO Special Programme for Research and Training in Tropical Diseases, TDR.

### Appendix: Open call strategies

| Ranking | Mean Score | Country | URL | URL (for non- English strategies) |
| --- | --- | --- | --- | --- |
| 1 | 44 | Peru and Columbia | https://docs.google.com/document/d/1gw0QUJGA59Ez2XK3YrKYBhapyE9HjPqVZRY51UR5fll/edit?usp=sharing | https://docs.google.com/document/d/1o_tVSLfHigFnptGJ6nSiyz33jTi_-jv688qVxFgT9vI/edit?usp=sharing |
| 2 | 41.73 | Nigeria | https://docs.google.com/document/d/1HoByeZT6wzHO8MGOavTrcKheSNJbTgLaUG5MjsTsx0/edit?usp=sharing | - |
| 3 | 39.67 | Uganda | https://docs.google.com/document/d/1extU9AKYODO2jrGtKX4siUdBO7tBfoWP/edit?usp=sharing&ouid=102190203285020963446&rtpof=true&sd=true | - |
| 4 | 39.25 | Nigeria | https://docs.google.com/document/d/15txDKWWj25Uwq_E3F83JR6ZeCRaVlExb/edit?usp=sharing&ouid=102190203285020963446&rtpof=true&sd=true | - |
| 5 | 38.36 | Uganda | https://drive.google.com/u/0/open?usp=forms_web&id=1pvi1rThdVgW6NM14Q0ZWTCaPpbOliX | - |
| 6 | 38.33 | Nigeria | https://docs.google.com/document/d/15d9No-T7J57ocQArEsWFTMob2yoarhbK/edit?usp=sharing&ouid=102190203285020963446&rtpof=true&sd=true | - |
| 7 | 37.63 | Bangladesh | https://docs.google.com/document/d/1D5t0FPLvjwuuMw6Rzyq4KGOzqj1S6twxA_w4X8gX3_s/edit?usp=sharing | - |
| 8 | 36.63 | China, Ethiopia | https://docs.google.com/document/d/1wRtAR6nei4ZEr6haldhklgjTetuG3wAb/edit?usp=sharing&ouid=102190203285020963446&rtpof=true&sd=true | - |
| 9 | 36.33 | Ethiopia | https://docs.google.com/document/d/1TIHClk8zzNKgHYTcoOXwaqG1Q8FVqBWk4RGbCBfTSmg/edit?usp=sharing | - |
| 10 | 35.83 | Ethiopia | https://docs.google.com/document/d/118mfYTBNVwgW7u10cLNjuYwtXlham0O4wiHj5puDtFU/edit?usp=sharing | - |
| 11 | 35.63 | Nigeria | https://docs.google.com/document/d/1FLqg7CMnX2xHaT3sDBLZUU0nfuXV3paA/edit?usp=sharing&ouid=102190203285020963446&rtpof=true&sd=true | - |
| 12 | 35.25 | India and other SEA LMIC | https://docs.google.com/document/d/1HO7rhcaHfdNz52z9ZsAWFfrvLI5JlhDJ/edit?usp=sharing&ouid=102190203285020963446&rtpof=true&sd=true | - |
| 13 | 34.33 | Peru and Columbia | https://drive.google.com/u/0/open?usp=forms_web&id=1mbWVdQIdRAxtmEW25T76s4Liz2KqYJfx | https://docs.google.com/document/d/1EzE3Z9--1rDhlYW-4RZCpzVg735AZO-IOOBX4LpWdK4/edit?usp=sharing |
| 14 | 34.33 | Yemen | https://docs.google.com/document/d/1Dua27_nwWNrX5QL9mcyxV5ZbrTkHKf1r/edit?usp=sharing&ouid=102190203285020963446&rtpof=true&sd=true | - |
| 15 | 34 | Nigeria | https://drive.google.com/u/0/open?usp=forms_web&id=18YZTPjLouiCH3uA6r2xWfdmctPT1B9zm | - |
| 16 | 33.93 | Nigeria | https://docs.google.com/document/d/1MO288R7PePm0Bcl0qJfPAVxF3JaXztEbWYmasy2W0Y/edit?usp=sharing | - |
| 17 | 32.67 | Columbia | https://drive.google.com/u/0/open?usp=forms_web&id=1_j_xzHLnXXcOxCNg9L5LiMIRvuD8TABl | https://docs.google.com/document/d/1W8ZTJ7pgTkrpFdBdieMNSuWkI6_pyJLum_tfS540Vi8/edit?usp=sharing |
| 18 | 32.33 | Ethiopia | https://drive.google.com/u/0/open?usp=forms_web&id=1KUrc4JLfXW55Z7mqt5JKPU2v_gveurAe | - |
| 19 | 32.33 | Argentina | https://drive.google.com/u/0/open?usp=forms_web&id=1BBctOgBC6YC0cP9Hgqjyp2rAL12NSjFE | https://docs.google.com/document/d/1W6hAWHG-GakyjekKZ_Nx9h25s6HTx5gDYDW8y4GhNXc/edit?usp=sharing |
| 20 | 32.33 | Nigeria | https://drive.google.com/file/d/1i32X0CplFXZVtEu1MBSfZmcOasXyYvWG/view | - |
| 21 | 32 | Rwanda and Guinea | https://drive.google.com/u/0/open?usp=forms_web&id=19gBoHtYnIMzQPee1mh2Z78S0zDTYWz4q | - |
| 22 | 32 | Malawi | https://drive.google.com/u/0/open?usp=forms_web&id=1xhgAzgHay_vY_DIP_H46o17gcoC6AI-9 | - |
| 23 | 31.33 | Nepal, Indonesia, Bangladesh | https://drive.google.com/u/0/open?usp=forms_web&id=1EQxj5iKpuAM5JBVOB4gvwQ35mn9cfpsX | - |
| 24 | 31 | Ethiopia | https://drive.google.com/u/0/open?usp=forms_web&id=1AShY5KPLJEwuHJhWF-P6UIW86OQLfZiU | - |
| 25 | 30.88 | N/A | https://docs.google.com/document/d/11H-uxID2UYLaJevdrZGGS2n7eCDdQvVPHMPYZkMp1PM/edit?usp=sharing | - |
| 26 | 29.63 | Nepal | https://docs.google.com/document/d/1whwdbmCT68pP0uR_8bjv8aRxyv0zKL1t/edit?usp=sharing&ouid=102190203285020963446&rtpof=true&sd=true | - |
| 27 | 29.6 | Ghana | https://drive.google.com/u/0/open?usp=forms_web&id=1Nde1VeVOTtoF1lzbRjhiROg-gCTvEgrG | - |
| 28 | 29.5 | Ethiopia and other LMICs | https://docs.google.com/document/d/1XKdX3ZB4LsQtQ3QU6YRsLBW_d74nU0axSDn21bICvLw/edit?usp=sharing | - |
| 29 | 29.33 | Columbia | https://drive.google.com/u/0/open?usp=forms_web&id=1fmII14YiNQFzDAsH2bsivz9VxQGx8Qlk | https://docs.google.com/document/d/1fbMtKZTyAGxIr2KHfYpk67TISO5pmYEEFSLWOFdXpbY/edit?usp=sharing |
| 30 | 29.25 | Tanzania | https://drive.google.com/u/0/open?usp=forms_web&id=1LBlQmEJwmJbQnvGUfQqhN1Lq-gOEEBRg | - |
| 31 | 29.05 | Several | https://docs.google.com/document/d/13Pkf7IXgPPUiztxlV86Lo_r_wD57Y58Cydeqzhqzu6c/edit?usp=sharing | - |
| 32 | 29 | Nigeria | https://drive.google.com/u/0/open?usp=forms_web&id=1L-6pm6mYMJ4RkCo-NiTzRxc2LTypdcGz | - |
| 33 | 28.62 | Bangladesh | https://drive.google.com/u/0/open?usp=forms_web&id=1mcuXW2kuWRyoRmLjCMsfXFk76xYfFLJU | - |
| 34 | 28.63 | Ethiopia | https://drive.google.com/u/0/open?usp=forms_web&id=1ryD-sjomip3kH4bSi6w4YpjutNVm6J7a | - |
| 35 | 28.43 | N/A | https://docs.google.com/document/d/1973NM4UFwEqNW52yURlhK8doL_-lgyp2/edit?usp=sharing&ouid=102190203285020963446&rtpof=true&sd=true | - |
| 36 | 28 | Nigeria | https://docs.google.com/document/d/1m2SGbOOQVuLWfiR8da_oYP2h4yaz7IZop/edit?usp=sharing&ouid=102190203285020963446&rtpof=true&sd=true | - |
| 37 | 27 | South Africa | https://docs.google.com/document/d/12BRrXACEQ3EVoJjwg49hvKGFbzagTbKE/edit?usp=sharing&ouid=102190203285020963446&rtpof=true&sd=true | - |
| 38 | 26.5 | USA | https://docs.google.com/document/d/1-vLsFzGqvxdSgJON6qSIp5fOkAnwMLu/edit?usp=sharing&ouid=102190203285020963446&rtpof=true&sd=true | - |
| 39 | 26.5 | Nigeria | https://drive.google.com/u/0/open?usp=forms_web&id=1BKXbPvS_ChIKWtlnM_ItbyHF8rGVgyRM | - |
| 40 | 25 | Kenya | https://drive.google.com/u/0/open?usp=forms_web&id=1_c56zCVDled4E6NhuLrNqzuTX9bWr9td | - |
| 41 | 24.5 | N/A | https://drive.google.com/u/0/open?usp=forms_web&id=1iwhUJvE4b6KgKOscmLzaCDs76Cnx6Cu4 | - |
| 42 | 23.5 | Yemen | https://drive.google.com/u/0/open?usp=forms_web&id=13M6FIYuX7r_ZFH-dCE_AXStncquKbjv | - |
| 43 | 23 | Malawi | https://drive.google.com/u/0/open?usp=forms_web&id=1d-tuKS_qwaVIIQSdUPJX89PLPyI4BKtM | - |
| 44 | 21.25 | 15 LMICs across the world | https://drive.google.com/u/0/open?usp=forms_web&id=1MuH1q6GSUSZVPmojespZT8oC0ft-zkzQ | - |
| 45 | 20 | N/A | https://drive.google.com/u/0/open?usp=forms_web&id=1UqOWfQLFP8zIWA89Mz9smku1F1SRbIKb | https://docs.google.com/document/d/1uogyjhhfeEx4Cy1r_3XyOsuCb81KLwguX3BtiSKK5OA/edit?usp=sharing |
| 46 | 16.25 | Malawi | https://drive.google.com/u/0/open?usp=forms_web&id=1gi_VaoHV6qJnknim1xZUHWuyirWc6Hlk | - |

